# symptomcheckR: an R package for analyzing and visualizing symptom checker performance

**DOI:** 10.1101/2024.02.06.24302384

**Authors:** Marvin Kopka, Markus A. Feufel

## Abstract

**Background:** A major stream of research on symptom checkers aims at evaluating the technology’s *predictive accuracy*, but apart from general trends, the results are marked by high variability. Several authors suggest that this variability might in part be due to different assessment methods and a lack of standardization. To improve the reliability of symptom checker evaluation studies, several approaches have been suggested, including standardizing input procedures, the generation of test vignettes, and the assignment of gold standard solutions for these vignettes. Recently, we suggested a third approach––test-theoretic metrics for standardized performance reporting–– to allow systematic and comprehensive comparisons of symptom checker performance. However, calculating these metrics is time-consuming and error prone, which could hamper the use and effectiveness of these metrics.

**Results:** We developed the R package symptomcheckR as an open-source software to assist researchers in calculating standard metrics to evaluate symptom checker performance individually and comparatively and produce publication-ready figures. These metrics include accuracy (by triage level), safety of advice (i.e., rate of correct or overtriage), comprehensiveness (i.e., how many cases could be entered or were assessed), inclination to overtriage (i.e., how risk-averse a symptom checker is) and a capability comparison score (i.e., a score correcting for case difficulty and comprehensiveness that enables a fair and reliable comparison of different symptom checkers). Each metric can be obtained using a single command and visualized with another command. For the analysis of individual or the comparison of multiple symptom checkers, single commands can be used to produce a comprehensive performance profile that complements the standard focus on accuracy with additional metrics that reveal strengths and weaknesses of symptom checkers.

**Conclusions:** Our package supports ongoing efforts to improve the quality of vignette-based symptom checker evaluation studies by means of standardized methods. Specifically, with our package, adhering to reporting standards and metrics becomes easier, simple, and time efficient. Ultimately, this may help users gain a more systematic understanding of the strengths and limitations of symptom checkers for different use cases (e.g., all-purpose symptom checkers for general medicine versus symptom checkers that aim at improving triage in emergency departments), which can improve patient safety and resource allocation.

## 1 Background

Symptom checkers––systems in which laypeople input symptoms to receive potential diagnoses and triage advice (1)–– are gaining popularity and drawing attention among health professionals and lay users as well as in the research community (2–4). Research in this field focuses, on one hand, on the potential impact of symptom checkers on individual users and healthcare systems (5–13), and on the accuracy of these systems on the other (2,3,14–17). For users, it is crucial that symptom checkers give safe advice and prevent potential harm (18), whereas for healthcare systems overtriage could inflate costs and strain scarce resources (e.g., due to unnecessary emergency department visits) (5,14). Thus, accurate performance is of great importance for symptom checker success, emphasizing the need of high standards for performance evaluations.

Evaluation studies of symptom checkers show high variability, however, with average accuracy estimates ranging from 27% to 90% (2,3). The reasons for the wide range of accuracy estimates are not entirely clear, although a first set of factors might relate to the choice of different evaluation methods, including testing procedures (19), types of case vignettes tested (19–21), and the gold standard solutions assigned to these cases (21,22). For instance, not all symptom checkers may be tested with every vignette (20) as some symptom checkers are designed for specialized tasks (such as only addressing pediatric cases), whereas other symptom checkers restrict the types of symptoms that may be entered and processed (16). As a result, accuracy cannot be effectively compared between these symptom checkers. A second factor might relate to the evaluation metrics used. For instance, to account for different goals such as avoiding individual harm and avoiding unnecessary demand on healthcare systems, some studies report additional metrics such as the safety of advice, although the exact metrics reported differ between studies (14,16,23,24).

As a remedy researchers proposed solutions to standardize evaluation methodologies: Painter et al. proposed several requirements, including standardizing the number of inputters, developing a standardized way of determining a gold standard solution to a case, or developing more reliable vignettes that are more representative for real-world cases (21). El-Osta et al. examined variability in the vignette creation processes and urged the field to use real-world data instead of artificial vignettes (22). Meczner et al. examined inputter variability and proposed that coding a vignette as solved (in)correctly by a symptom checker should involve multiple coders and a synthesis of their assessments (25).

In a more recent study, our research team proposed guidelines to enhance symptom checker *reporting*. The metrics we developed focus solely on triage accuracy (as opposed to diagnostic accuracy), because the (final) diagnosis is invariably made by a healthcare professional and is thus a less relevant feature of symptom checkers (16,17,26). The guidelines include various metrics that provide insights into individual symptom checker performance as well as performance comparisons (20). Most importantly, we suggested quality indicators to control for bias in comparative accuracy estimates (e.g., such as how many cases a symptom checker could be tested with) and to guide the selection of symptom checkers for a specific use case (e.g., implementing it in an emergency department or using it for at-home testing). Using these quality indicators, comparability across different studies can be enhanced and implementation can be guided in a manner that is specific to each use case. To account for sources of bias in the accuracy estimate, we developed a ‘Capability Comparison Score’ based on classic test theory that adjusts for the difficulty and number of cases entered to allow reliable comparisons between different symptom checkers.

Although most of the proposed metrics can be calculated easily, it is neither cost efficient nor practical for researchers to calculate all metrics by hand. To solve this problem, software solutions can be used. Currently, there is only the psych package (27) available, which can calculate item difficulty, but no other metrics specific for symptom checker evaluations. Since no software is available to assess the performance of one symptom checker or a comparison of multiple ones in a standardized way, future studies are likely to continue reporting differing metrics, which limits the comparability between them. To improve quality standards in symptom checker research, we developed an R package named *symptomcheckR*, which assists users in calculating and reporting standardized metrics on symptom checker performance.

## 2 Implementation

The R-Package was developed to include several metrics for evaluating the (comparative) performance of symptom checkers using the data of the abovementioned publication as case study (20). The metrics complement the commonly used single accuracy measure by shedding light both on its strengths *and* weaknesses. The package is optimized for easeof-use to allow symptom checker researchers, developers, policymakers, and other stakeholders to quickly analyze the performance of single or multiple symptom checkers. To achieve this, we adhered to key usability principles, ensuring the software is (a) effective by enabling users to generate comprehensive metrics, (b) efficient through providing single commands for each outcome and a unified command structure for all metrics, and (c) easy to use by including an example dataset to simplify data wrangling and commands inspired by natural language (28,29). The package builds on the previously published packages dplyr (30), tidyr (31), ggplot2 (32) and ggpubr (33). It is available on CRAN with open-source code and licensed under the GNU General Public License.

### 2.1 Metrics

In this section, we describe the developed metrics. The first five metrics are designed to evaluate the performance of an individual symptom checker, but they may be used to compare different symptom checkers as well. What is necessary for a comparative analysis is the focus of the subsequent metrics: item difficulty can be used to assess the difficulty of vignettes across symptom checkers, whereas the capability comparison score serves as a metric to account for potential sources of bias that may affect accuracy (e.g., how many easy and difficult vignettes could be entered). This ensures more reliable comparisons of the capabilities of different symptom checkers.

#### 2.1.1 Accuracy

Accuracy is defined as the proportion of cases that a symptom checker successfully solves. Although this metric provides an initial insight into the performance of an individual symptom checker, it does not account for varying levels of case urgency or the difficulty of the cases. It can be calculated as:

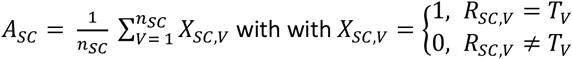

where A denotes the accuracy, n_SC_ the number of cases a symptom checker was tested with, V the vignette’s number, X_SC,V_ whether a case was solved correctly (with R_SC, V_ denoting the recommendation’s triage level and T_V_ the correct triage level).

#### 2.1.2 Accuracy by triage level

To gain more comprehensive insights on how symptom checkers perform in different scenarios, accuracy should be calculated for each triage level separately. For example, some symptom checkers do not advice self-care (16) and are thus not suitable to use on such cases. However, this information is not inferable from an aggregate accuracy. Another example is a symptom checker for emergency departments, which should distinguish particularly well between emergency and non-emergency cases. The following metric thus examines the use-case specific accuracy. The accuracy for each triage level can be calculated as:

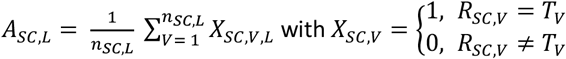

where A_SC,L_ denotes the accuracy the accuracy for a symptom checker on triage level L, n_SC,L_ the number of cases a symptom checker was tested with on the triage level L, V the vignette’s number, and X_SC,V,L_ whether a case was solved correctly (with R_SC,V_ denoting the recommendation’s triage level and T_V_ the correct triage level).

#### 2.1.3 Safety of advice

The safety of advice gives an impression on how safe recommendations by a symptom checker are. This might be particularly relevant when evaluating the potential harm of a symptom checker. It indicates the percentage of recommendations that are categorized as being of equal or greater urgency than what is appropriate for a given case and can be calculated as:

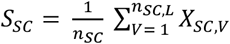

with 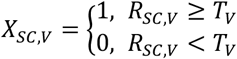

where S_SC_ denotes the safety, n_SC_ the number of cases a symptom checker was tested with, V the vignette’s number, and X_SC,V_ whether the recommendation of a symptom checker SC for a vignette V was safe (with R denoting the recommendation’s triage level –higher values indicating higher urgency – and T the correct triage level).

#### 2.1.4 Comprehensiveness

Not all symptom checkers allow entering all cases. If only few symptoms can be entered, a symptom checker might be beneficial for a specific use case, but not for broad implementation. Further, entering only selected cases can bias the accuracy. Thus, the comprehensiveness metric accounts for how many cases could be entered in a symptom checker and can be calculated as:

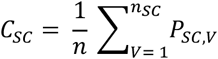

where C_SC_ denotes the comprehensiveness, n the total number of vignettes in the set, and P_SC,V_ whether a symptom checker provided a recommendation for this vignette.

#### 2.1.5 Inclination to overtriage

Whereas providing safe advice is essential to protect individuals from harm, frequently giving advice with (unnecessarily) high urgency can result in increased healthcare utilization due to the use of a symptom checker (34). This, in turn, can lead to increased healthcare expenditures and reduced availability of care resources for individuals (5,10). Thus, assessing a symptom checker’s inclination to overtriage is especially valuable from a systems perspective and can be quantified as the proportion of ‘overtriage’ errors among all incorrect triage recommendations. It can be calculated as:

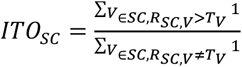

where ITO_SC_ denotes the symptom checker’s inclination to overtriage, V the vignette, SC the symptom checker, R_SC,V_ the recommendation of a symptom checker SC for the vignette V and T_V_ the correct triage level for the vignette V.

#### 2.1.5 Item Difficulty

When testing multiple symptom checkers with the same cases, some cases might be solved by all symptom checkers and some by none. Item difficulty can be used to determine how difficult a vignette is for symptom checkers to solve. It describes the proportion of symptom checkers that were able to solve a vignette – thus, an item difficulty of 1 means that the case was easy to solve (as all symptom checkers solved it) and an item difficulty of 0 means that it is particularly difficult (as none solved it correctly). It can be calculated as:

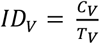

where ID denotes the item difficulty, C_V_ the number of symptom checkers that solved a vignette correctly and T_V_ the total number of symptom checkers that assessed the case.

#### 2.1.5 Capability Comparison Score

Because not all symptom checkers can be tested with all cases and those cases that can be entered differ in their difficulty (20), solely comparing different symptom checkers according to their accuracy results in biased conclusions. Thus, the capability comparison score accounts for the fact that (a) not all symptom checkers are tested with the same cases and (b) these cases differ in difficulty. It allows more reliable performance comparisons between different symptom checkers and can be calculated as:

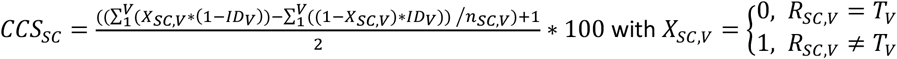

where CCS denotes the resulting score, SC the symptom checker that is being assessed, X_SC,V_ whether the advice was correct (with R denoting the recommendation’s triage level – higher values indicating higher urgency – and T the correct triage level), ID the item difficulty, V the vignette, and n_SC,V_ the number of cases that were entered in the symptom checker.

### 2.2 Visualization

The symptomcheckR can be used to create publication-ready stacked bar plots to visualize all metrics for individual symptom checkers: accuracy (by triage level), safety of advice, comprehensiveness, and inclination to overtriage. Additionally, it can be used to create double-sided bar charts for illustrating the capability comparison score. All charts are color coded for intuitive understanding: desirable outcomes are shown in green, while undesirable ones are red. The color shades are chosen in accordance with inclusive design standards, ensuring they are distinguishable to individuals with color vision deficiencies. The package also includes two additional commands: one to visualize the performance of a single symptom checker across all metrics, and another for a side-by-side performance comparison of multiple symptom checkers. These commands return a ggplot class object, which can be further customized to meet various design requirements and preferences. For combined performance visualizations, the command returns a ggarrange class object.

### 2.3 Included Dataset

In the R package, we included a dataset derived from a previous study on the accuracy of different symptom checkers (16,35). This study tested different freely accessible symptom checkers in 2020 using a set of 45 vignettes, initially developed by Semigran et al. (17). It comprises several symptom checkers with varying degrees of comprehensiveness and can thus be used as an example dataset to demonstrate the different functions of the package.

## 3 Results

To demonstrate the usage of the symptomcheckR package, we conduct a full analysis of the included dataset using all commands available in the package. This analysis conforms to the reporting standards recommended for symptom checker audit studies (20). We present the analysis of both evaluating a single symptom checker and comparing multiple symptom checkers.

### 3.1 Dataset

The dataset can be loaded using the *data(symptomcheckRdat*a) command. It comprises 22 symptom checkers which were tested with 45 vignettes each, yielding a total sample size of 990 observations. 19.6% (194/990) are missing data, i.e. include cases in which a symptom checker did not provide a recommendation.

## 3.2 Analysis of individual symptom checker performance

As an individual symptom checker, we selected Ask NHS, because it contains missing data and can be used to show the possibilities of all commands. The first step is analyzing its accuracy. This can be done using the *get_accuracy()* command. It includes the arguments *data* for specifying the dataset and *correct* (as a string) to indicate the column in which correct responses are stored as a Boolean (TRUE or FALSE). It returns a single accuracy value:

~~~
*accuracy
1 0.6060606*
~~~

This result can be visualized using *plot_accuracy()*, see Figure 1. The following exemplary code first obtains a new data frame containing only ASK NHS data and then shows the analysis using base R and using dplyr:

**Figure 1.**
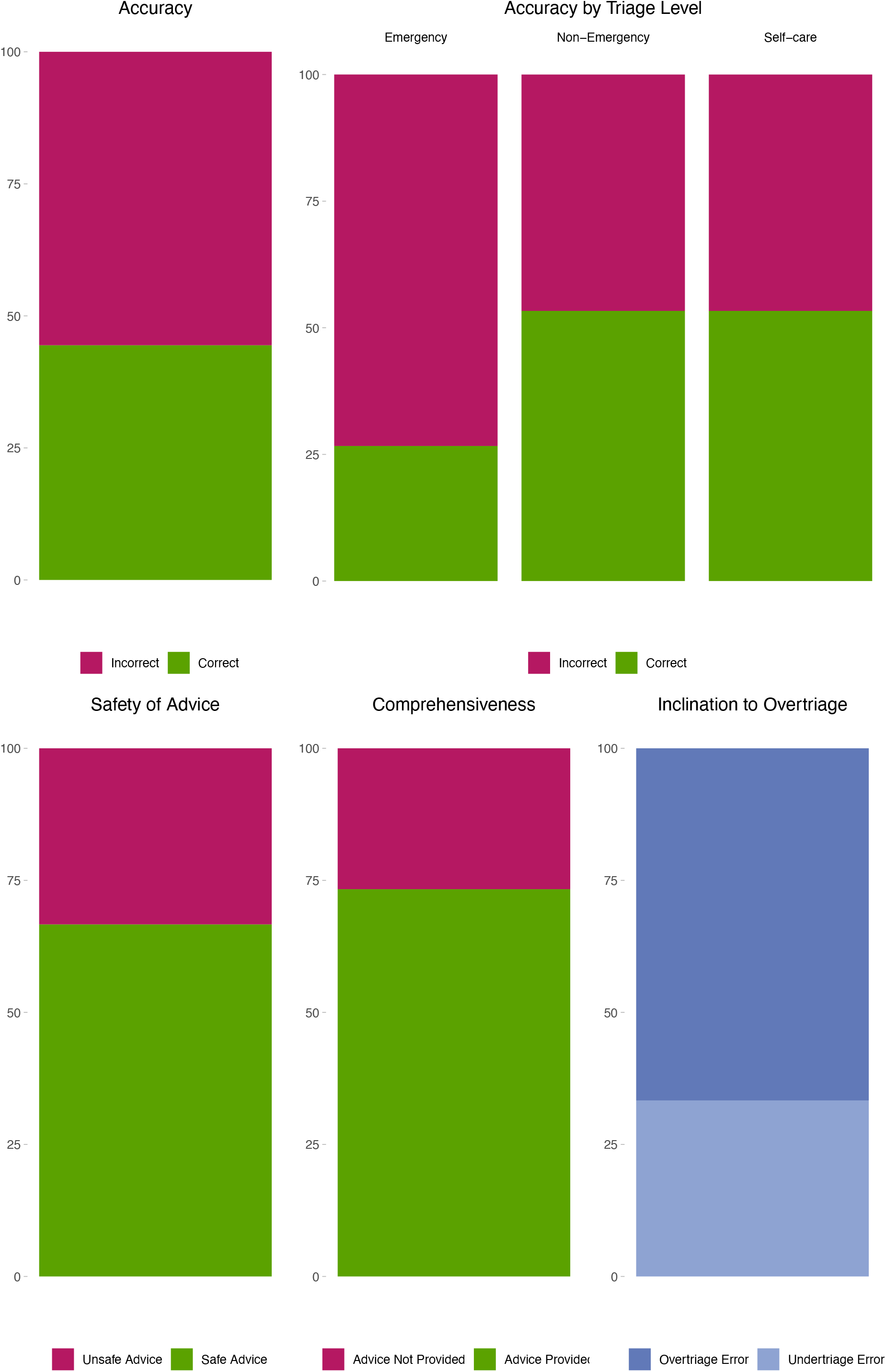
Publication-ready figure from using the plot_performance_single() function. The figure visualizes all relevant metrics for a symptom checker’s triage performance (in this case Ask NHS).

~~~
*data(symptomcheckRdata)
df_individual <-symptomcheckRdata %>%
 filter(App_name == “Ask NHS”)
# Using base R
accuracy_value <-get_accuracy(df_individual, correct = “Correct_Triage_Advice_provided_from_app”)
plot_accuracy(accuracy_value)
# Using dplyr
df %>%
get_accuracy(correct = “Correct_Triage_Advice_provided_from_app”) %>%
plot_accuracy()*
~~~

Next, the accuracy can be analyzed for each triage level using the *get_accuracy_by_triage()* function. It includes the same arguments as the *get_accuracy()* command and adds a *triagelevel* (as a string) argument denoting the column in which the correct triage solution is stored. The output can look like this:

~~~
*Goldstandard_solution accuracy
<chr> <dbl>
1 Emergency 0.333
2 Non-Emergency 0.727
3 Self-care 0.8*
~~~

This can again be visualized using *plot_accuracy_by_triage_level()*. The following code shows this process:

~~~
*accuracy_by_triage <-get_accuracy_by_triage(data = df_individual,
                     correct = “Correct_Triage_Advice_provided_from_app”,
                      triagelevel = “Goldstandard_solution”)
plot_accuracy_by_triage(accuracy_by_triage)*
~~~

Next, users can visualize the safety of the advice. This can be done using the *get_safety_of_advice()* command with the arguments *data* for specifying the dataset, *triagelevel_correct* (as a string) for specifying the column in which the correct triage level solutions are stored, triagelevel_advice (as a string) for specifying the column in which the symptom checker recommendations are stored, and *order_triagelevel* (as a vector) for specifying the order of triage levels, starting with the level of highest urgency. The output looks like this:

~~~
*$raw_numbers
# A tibble: 2 × 2
# Groups: safety [2]
safety n
<chr> <int>
1 safe advice 22
2 unsafe advice 11
$percentage
# A tibble: 1 × 1
 safety_percentage
   <dbl>
1 66.7*
~~~

It can be visualized using *plot_safety_of_advice()*, which takes the input of the first command again. The code looks like this:

~~~
*safety <-get_safety_of_advice(data = df_individual,
                          triagelevel_correct = “Goldstandard_solution”,
                          triagelevel_advice = “Triage_advice_from_app”,
                          order_triagelevel = c(“Emergency”, “Non-Emergency”, “Self-care”))
plot_safety_of_advice(safety)*
~~~

Afterwards, the comprehensiveness can be of interest. It can be calculated with the command *get_comprehensiveness()* with the arguments *data* for specifying the dataset, *triagelevel_advice* (as a string) for specifying the column in which the symptom checker recommendations are stored, and *vector_not_entered* (as a vector) for specifying all values that are coded as no recommendation from a symptom checker or no possibility to enter the case. The output looks like this:

~~~
*$raw_numbers
     # A tibble: 2 × 2
     # Groups: gave_advice [2]
         gave_advice n
        <chr> <int>
       1 Advice given 33
       2 Advice not given 12
       $percentage
       # A tibble: 1 × 1
         comprehensiveness_percentage
                       <dbl>
       1                73.3*
~~~

It can again be visualized using *plot_comprehensiveness()* with the result of *get_comprehensiveness()* as the input. The code looks as follows:

~~~
*comprehensiveness <-get_comprehensiveness(data = df_individual,
                                  triagelevel_advice = “Triage_advice_from_app”,
                                  vector_not_entered = c(NA))
plot_comprehensiveness(compreheniveness)*
~~~

Lastly, the inclination to overtriage can be calculated using *get_inclination_overtriage()* with the arguments *data* for spec-ifying the dataset, *triagelevel_correct* (as a string) for specifying the column in which the correct triage level solutions are stored, *triagelevel_advice* (as a string) for specifying the column in which the symptom checker recommendations are stored, and *order_triagelevel* (as a vector) for specifying the order of triage levels. The triage levels are sorted by urgency, starting with the highest urgency first and the lowest urgency last. The output looks like this:

~~~
*$raw_numbers
        # A tibble: 2 × 2
        # Groups: overtriage_undertriage [2]
           overtriage_undertriage n
           <chr> <int>
        1 overtriage 11
        2 undertriage 22
        $percentage
        # A tibble: 1 × 1
          inclination_to_overtriage_percentage
                                <dbl>
          1                33.3*
~~~

It can be visualized using plot_inclination_overtriage(). The code looks like the following:

~~~
*inclination_to_overtriage <-get_inclination_overtriage(data = df_individual,
                triagelevel_correct = “Goldstandard_solution”,
                triagelevel_advice = “Triage_advice_from_app”
                order_triagelevel = c(“Emergency”, “Non-Emergency”, “Self-care”))
plot_inclination_overtriage(inclination_to_overtriage)*
~~~

To get a comprehensive overview of a symptom checker’s performance, all metrics can be visualized in a single plot using *plot_performance_single()* with the arguments *data* for specifying the dataset, *triagelevel_correct* (as a string) for specifying the column in which the correct triage level solutions are stored, triagelevel_advice (as a string) for specifying the column in which the symptom checker recommendations are stored, *order_triagelevel* (as a vector) for specifying the order of triage levels, and *vector_not_entered* (as a vector) for specifying all values that are coded as no recommendation from a symptom checker or no possibility to enter the case. The code can look like this:

~~~
plot_performance_single(data = df_individual,
     triagelevel_correct = “Goldstandard_solution”,
     triagelevel_advice = “Triage_advice_from_app”,
     order_triagelevel = c(“Emergency”, “Non-Emergency”, “Self-care”),
     vector_not_entered = c(NA))
~~~

The resulting figure can be seen in Figure 1.

## 3.2 Performance comparison of multiple symptom checkers

The same commands can be used to compare multiple symptom checkers. To change the functions’ output to comprise multiple symptom checkers, the *apps* argument (as a string) can be added to indicate the column in which the names of different symptom checkers are stored. A full analysis with the same commands as those employed for the evaluation of individual symptom checkers could be conducted as follows:

~~~
*accuracy_value <-get_accuracy(symptomcheckRdata, correct = “Correct_Triage_Advice_provided_from_app”,
                               apps = “App_name”)
plot_accuracy(accuracy_value)
get_accuracy_by_triage(symptomcheckRdata,
                     correct = “Correct_Triage_Advice_provided_from_app”,
                     triagelevel = “Goldstandard_solution”,
                     apps = “App_name”)
plot_accuracy_by_triage(accuracy_value_by_triage)
safety <-get_safety_of_advice(data = symptomcheckRdata,
                         triagelevel_correct = “Goldstandard_solution”,
                         triagelevel_advice = “Triage_advice_from_app”,
                         order_triagelevel = c(“Emergency”, “Non-Emergency”, “Self-care”),
                         apps = “App_name”)
plot_safety_of_advice(safety)
comprehensiveness <-get_comprehensiveness(data = symptomcheckRdata,
                          triagelevel_advice = “Triage_advice_from_app”,
                          vector_not_entered = c(NA),
                          apps = “App_name”)
plot_comprehensiveness(comprehensiveness)
inclination_to_overtriage <-get_inclination_overtriage(data = symptomcheckRdata,
                                triagelevel_correct = “Goldstandard_solution”,
                                triagelevel_advice = “Triage_advice_from_app”,
                                order_triagelevel = c(“Emergency”, “Non-Emergency”, “Self-care”),
                                apps = “App_name”)
plot_inclination_overtriage(inclination_to_overtriage)*
~~~

Additionally, users can calculate the item difficulty and a capability comparison score to compare different symptom checkers. The item difficulty can be obtained using *get_item_difficulty()* with the arguments *data* for specifying the dataset, *correct* (as a string) to indicate the column in which correct responses are stored as a Boolean (TRUE or FALSE), and *vignettes* (as a string) to indicate the column in which the vignettes (as numbers or characters) are stored. The capability comparison score can be calculated using *get_ccs()* with the same arguments and an *apps* (as a string) argument indicating the column in which different symptom checker names are stored. It can also be calculated for different triage levels using *get_ccs_by_triage()* with the additional argument *triagelevel* (as a string) to indicate the column in which the correct triage level solutions are stored. Both can be visualized using plot_ccs() and plot_ccs_by_triage(), see Figure 2. An exemplary code can look like this:

**Figure 2.**
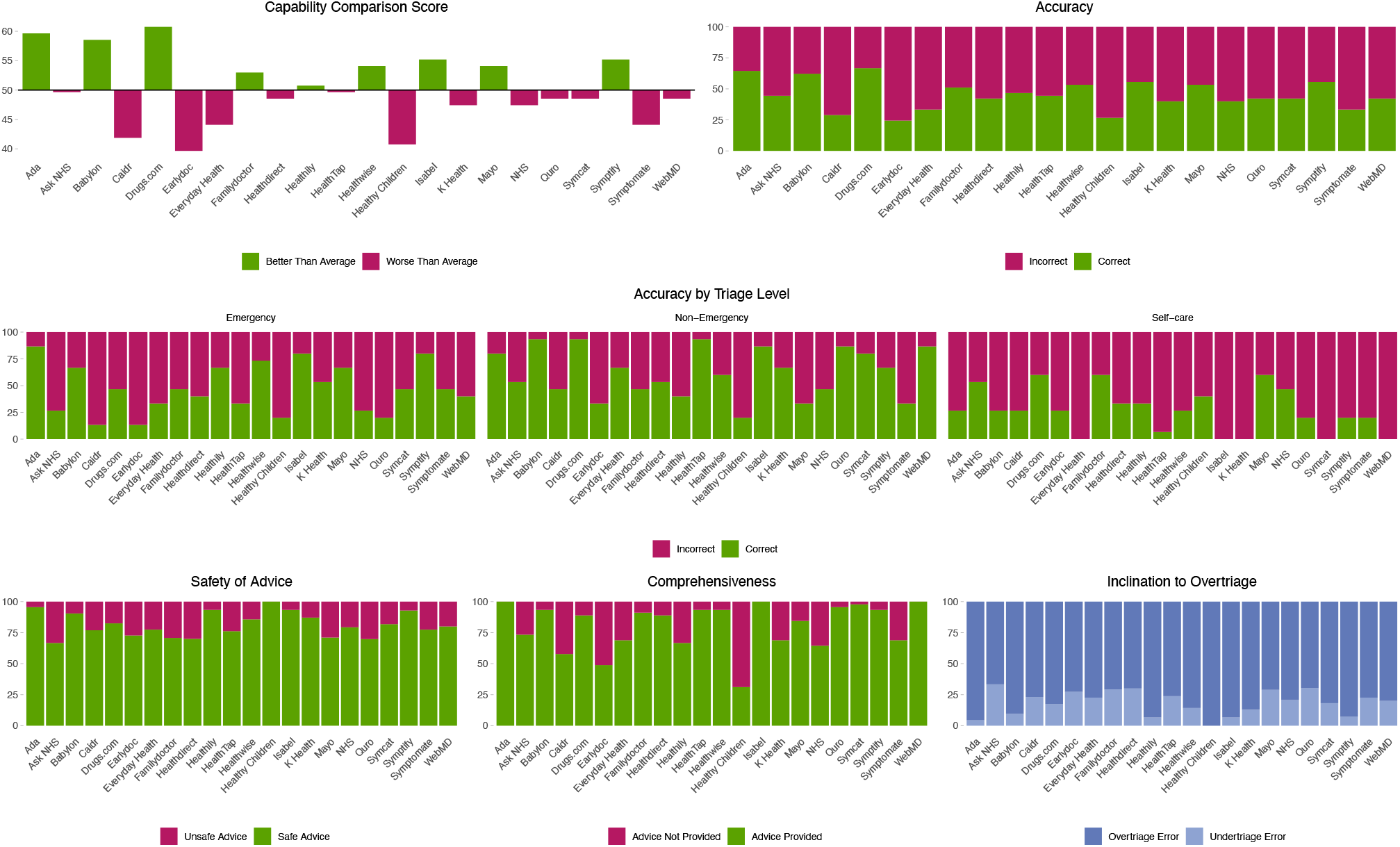
Publication-ready figure from using the plot_performance_multiple() function. The figure visualizes all relevant metrics for comparing different symptom checkers’ triage performance.

~~~
*get_item_difficulty(data = symptomcheckRdata,
                 correct = “Correct_Triage_Advice_provided_from_app”,
                 vignettes = “Vignette_id”)
ccs <-get_ccs(data = symptomcheckRdata,
         correct = “Correct_Triage_Advice_provided_from_app”,
         vignettes = “Vignette_id”,
         apps = “App_name”)
plot_ccs(ccs)
get_ccs_by_triage <-get_ccs_by_triage(data = symptomcheckRdata,
                         correct = “Correct_Triage_Advice_provided_from_app”,
                         vignettes = “Vignette_id”,
                         apps = “App_name”,
                        triage = “Goldstandard_solution”)*
~~~

Finally, the full dataset can be analyzed and visualized with the command *plot_performance_multiple()* with the arguments *data* for specifying the dataset, *triagelevel_correct* (as a string) for specifying the column in which the correct triage level solutions are stored, triagelevel_advice (as a string) for specifying the column in which the symptom checker recommendations are stored, *order_triagelevel* (as a vector) for specifying the order of triage levels, *vector_not_entered* (as a vector) for specifying all values that are coded as no recommendation from a symptom checker or no possibility to enter the case, *vignettes* (as a string) to indicate the column in which the vignettes (as numbers or characters) are stored, and *apps* (as a string) to indicate the column in which the names of different symptom checkers are stored. This results in a figure containing a comparison of all symptom checkers in the dataset across all metrics (see Figure 2). In this figure, the performance of all metrics is readily apparent. For instance, to identify a symptom checker suitable for general implementation, one should first examine the comprehensiveness section. This helps to rule out symptom checkers that are limited to entering certain cases only. Subsequently, the capability comparison scores can be examined to pinpoint symptom checkers that perform well. After narrowing down the choices, they can be evaluated with respect to their safety to ensure there is no potential harm to users. Additionally, examining the inclination to overtriage can be crucial to determine if it might unduly burden healthcare resources. This process can be repeated and adapted to various usecases, aiding in the selecting the most appropriate symptom checker for a specific use-case. The code to obtain such a figure can look like this:

~~~
*plot_performance_multiple(data = symptomcheckRdata,
                       triagelevel_correct = “Goldstandard_solution”,
                       triagelevel_advice = “Triage_advice_from_app”,
                       order_triagelevel = c(“Emergency”, “Non-Emergency”, “Self-care”),
                       vector_not_entered = c(NA)
                       vignettes = “Vignette_id”,
                       apps = “App_name”)*
~~~

## 4 Discussion

Whereas existing packages such as the psych package (27) offer item difficulty calculation but lack metrics specifically tailored to symptom checkers, the symptomcheckR package presented in this paper is designed to help analyze and visualize various performance metrics of individual symptom checkers and for performance comparisons. Whereas previous studies often focused solely on reporting accuracy, the metrics described reveal potential sources of bias in accuracy and allow drawing more reliable conclusions about the strengths and weaknesses of symptom checkers. For instance, as can be seen in Figure 2, WebMD had medium accuracy overall. However, a more detailed examination reveals that it is among the best-performing symptom checkers for identifying non-emergency care cases, yet one of the worst performing for self-care cases. WebMD also shows a high comprehensiveness, as all cases could be entered. Such nuances would remain hidden if the analysis were limited to accuracy and become apparent when examining the performance of symptom checkers in such a comparative figure. This way, our package contributes to ongoing research efforts aimed at standardizing evaluation methods and enhancing the quality of symptom checker assessments. There is an increasing number of studies offering recommendations or stating requirements to improve symptom checker assessments based on exploring the effect of different methodological variations (e.g., inputter instructions or gold standard solution assignment) (21,22). Because only few studies tend to implement these standards, we believe it is crucial to supplement empirical research with user-friendly software to facilitate implementation of these standards.

Our package has some limitations: it focuses mainly on triage accuracy and does not include commands for assessing diagnostic performance (e.g., evaluating the top diagnosis, the top three, or top twenty diagnoses (17)). However, our accuracy commands may be used for diagnostic accuracy by coding the corresponding responses in a new variable as true or false and using the *get_accuracy()* command. Secondly, users may have collected and stored their data in formats different from our example dataset. Because our package requires a specific data format, users will need to adjust their data format accordingly. To assist with this, we provide an example dataset to facilitate data wrangling. Lastly, the package incorporates current reporting standards. As new metrics emerge, they can be integrated into future versions of the package.

## 5 Conclusions

The symptomcheckR package is the first software that enables users to analyze the performance of symptom checkers using multiple metrics and produce publication-ready figures. It also allows more reliable comparisons of different symptom checkers, comprehensive insights into various aspects of their performance, and increases transparency in symptom checker audit studies. Consequently, users can determine the most appropriate symptom checker for a specific use case (e.g., integration in an emergency department) and identify factors that may influence accuracy estimates (such as the exclusive testing of simpler vignettes). These functionalities make the package especially useful for researchers, as well as for developers and regulatory bodies. We thus encourage these stakeholders to utilize the symptomcheckR package. If used widely, reliable, transparent, and easy-to-use evaluation and reporting standards may help to realize the potential of digital health innovations to improve patient safety and optimize the allocation of healthcare resources. We thus invite the community to contribute to improvements of the package and to develop their own software for other parts of symptom checker evaluation methodology.

## Data Availability

The datasets analyzed in the current study are available in the symptomcheckR package, available on CRAN.

https://cran.r-project.org/web/packages/symptomcheckR/index.html

